# Multiple introductions followed by ongoing community spread of SARS-CoV-2 at one of the largest metropolitan areas in the Northeast of Brazil

**DOI:** 10.1101/2020.08.25.20171595

**Authors:** Marcelo Henrique Santos Paiva, Duschinka Ribeiro Duarte Guedes, Cássia Docena, Matheus Filgueira Bezerra, Filipe Zimmer Dezordi, Laís Ceschini Machado, Larissa Krokovsky, Elisama Helvecio, Alexandre Freitas da Silva, Luydson Richardson Silva Vasconcelos, Antonio Mauro Rezende, Severino Jefferson Ribeiro da Silva, Kamila Gaudêncio da Silva Sales, Bruna Santos Lima Figueiredo de Sá, Derciliano Lopes da Cruz, Claudio Eduardo Cavalcanti, Armando de Menezes Neto, Caroline Targino Alves da Silva, Renata Pessôa Germano Mendes, Maria Almerice Lopes da Silva, Michelle da Silva Barros, Wheverton Ricardo Correia do Nascimento, Rodrigo Moraes Loyo Arcoverde, Luciane Caroline Albuquerque Bezerra, Sinval Pinto Brandão Filho, Constância Flávia Junqueira Ayres, Gabriel Luz Wallau

**Affiliations:** Núcleo de Ciências da Vida, Centro Acadêmico do Agreste, Universidade Federal de Pernambuco (UFPE), Universidade Federal de Pernambuco, Centro Acadêmico do Agreste - Rodovia BR-104, km 59 - Nova Caruaru, Caruaru - PE - Brasil. CEP: 55002-970; Instituto Aggeu Magalhães, Fundação Oswaldo Cruz, Departamento de Entomologia. Av. Professor Moraes Rego, s/n - Campus da UFPE - Cidade Universitária. Recife/PE – Brasil. CEP: 50.740-465; Instituto Aggeu Magalhães, Fundação Oswaldo Cruz, Núcleo de Plataformas Tecnológicas (NPT); Instituto Aggeu Magalhães, Fundação Oswaldo Cruz, Departamento de Microbiologia; Instituto Aggeu Magalhães, Fundação Oswaldo Cruz, Departamento de Parasitologia; Instituto Aggeu Magalhães, Fundação Oswaldo Cruz, Laboratório de Virologia e Terapia Experimental (LAVITE); Universidade Federal de Pernambuco (UFPE), Centro de Ciências Médicas, Departamento de Medicina Tropical. Av. Professor Moraes Rego, s/n - Campus da UFPE - Cidade Universitária. Recife/PE – Brasil; Secretaria de Saúde de Pernambuco (SES-PE), Secretaria Executiva de Vigilância em Saúde (SEVS-PE). Rua Dona Maria Nogueira, 519, Bongi. Recife/PE, Brasil; Instituto Aggeu Magalhães, Fundação Oswaldo Cruz, Núcleo de Bioinformática (NBI)

## Abstract

The emergence of SARS-CoV-2 in the human population has caused a huge pandemic that is still unfolding in many countries around the world. Multiple epicenters of the pandemic have emerged since the first pneumonia cases in Wuhan, first in Italy followed by the USA and Brazil. Up to now, Brazil is the second most affected country, however, genomic sequences of SARS-CoV-2 strains circulating in the country are restricted to some highly impacted states. Although the Pernambuco state, located in the Northeast Region, is the sixth most affected brazilian state and the second considering lethality rate, there is a lack of high quality genomic sequences from the strains circulating in this region. Here, we sequenced 38 strains of SARS-CoV-2 from patients presenting Covid-19 symptoms. Phylogenetic reconstructions revealed that three lineages were circulating in the state and 36 samples belong to B1.1 lineage. We detected two introductions from European countries and five clades, corroborating the community spread of the virus between different municipalities of the state. Finally, we detected that all except one strain showed the D614G spike protein amino acid change that may impact virus infectivity in human cells. Our study brought new light to the spread of SARS-CoV-2 strains in one of the most heavily impacted states of Brazil.

## INTRODUCTION

Severe Acute Respiratory Syndrome (SARS) is caused by several viral pathogens that are able to infect the upper and lower respiratory tract of humans leading to compromised oxygenation and multiple organ failure [1]. Influenza viruses, rhinoviruses and coronaviruses (CoV) are commonly associated with SARS, although with a low incidence due to the worldwide influenza immunization campaigns and the mild symptoms normally derived from infection of other respiratory viruses [2]. On the other hand, recently emerged respiratory viruses, which are able to sustain human-to-human transmission, may infect a large proportion of the world human population, leading to devastating pandemics in the absence of a vaccine and/or preventive measures to control its dissemination. For example, the H1N1 virus infected more than 575 thousand people during the 2009 pandemic [3] and two coronaviruses, SARS-CoV-1 that emerged in 2002 and MERS-CoV, in 2012, infected thousands of people. These last two viruses were not well adapted to human-to-human transmission and the outbreaks came into control before massive human spread [4]

Viruses from the Coronaviridae family have a positive sense RNA genome ranging from 27 to 32 Kb in length and codify 16 non-structural and 4 structural proteins [5]. These highly diverse viruses are composed by at least five genuses (Alphaletovirus, Alphacoronavirus, Betacoronavirus, Deltacoronavirus and Gamacoronavirus) that are infectious to a myriad of vertebrates [6,7]. Six different coronaviruses from Alpha and Betacoronavirus genuses are known to infect humans, all of them likely derived from wild and domestic animals [5,8]. Studies suggest that four of these viruses are associated with only mild symptoms in humans (HCoV-NL63, HCoV-229E, HCoV-OC43 and HKU1), while SARS-CoV-1 and MERS-CoV show a high morbidity and mortality rate (MASTERS, 2006; SINGHAL, 2020). However, around December 2019 a new pneumonia-like illness was reported in the Wuhan municipality in China (ZHOU et al., 2020). A few weeks later the genome of the etiological agent was sequenced and revealed a new coronavirus named SARS-CoV-2 [9]. Different from the SARS-CoV-1 and MERS-CoV, SARS-CoV-2 is particularly well adapted to sustained human-to-human transmission. It is highly infectious, spreading easily through symptomatic and asymptomatic individuals [10]. Due to widespread human mobility, this virus reached all continents in less than 3 months after the reports from Wuhan (https://www.who.int/emergencies/diseases/novel-coronavirus-2019/situation-reports/). Such a pandemic is still unfolding and more than 15 million people were already infected by the virus, and more than 600 thousand succumbed, highlighting that the human kind is facing the largest and more challenging human pandemic of all times.

Since the initial spread of SARS-CoV-2, there were two well recognized epicenters of the pandemic: Northern Italy that reported the peak of the epidemic around late March to mid April 2020 [11,12] followed by the United States of America (USA), which reached the first peak around April 2020, with most cases concentrating in the New York City area [13]. Currently, the USA is the most affected country in the world, accounting for almost five million infected people. Real time epidemiological monitoring are showing that the numbers of new infected patients are decreasing in some early affected states while increasing rapidly in later affected ones giving support to a second wave in the country [14] (https://coronavirus.jhu.edu/map.html, https://www.who.int/emergencies/diseases/novel-coronavirus-2019/situation-reports/). On the other hand, SARS-CoV-2 human infections have been increasing at a rapid pace in South America, led mostly by Brazil - the most populous country of the continent with more than 211.8 million people according to the Brazilian Institute of Geography and Statistics (Instituto Brasileiro de Geografia e Estatística, IBGE - http://www.ibge.gov.br). The first SARS-CoV-2 infection in Brazil was reported in February 25th 2020, and now (July 2020), Brazil is the second most affected country in the world, only behind the US in the number of new cases and deaths [15] (https://coronavirus.jhu.edu/map.html - https://covid.saude.gov.br/). Recent publications based on the sequencing and characterization of SARS-CoV-2 genomes from Minas Gerais and São Paulo (southeast region of Brazil) revealed the introduction of different SARS-CoV-2 lineages from European countries, which were associated with recent travel history of the patients [16-18]. Resende et al 2020 [19], sequenced 95 SARS-CoV-2 genomes from 10 brazilian states and Candido et al 2020 sequenced 427 genomes from 21 Brazilian states and both identified more than 100 independent introduction of SARS-CoV-2 in Brazil [19,20]. Pernambuco is the seventh most populous state in Brazil and the eighth state regarding the number of infected patients with 101.395 confirmed cases, but the fourth in terms of deaths (6.828 deaths) and the second considering lethality (6.7%) (last accessed August 8th 2020 - CIEVS PE - https://www.cievspe.com/). So far no genomic epidemiology study [21] was performed to sequence SARS-CoV-2 viral genomes circulating in the state.

In this study, we sequenced 38 SARS-CoV-2 genomes from the Pernambuco state, Northeastern Brazil during an early pandemic phase in order to evaluate the emergence and community spread of this virus in one of the most affected states. We found that two introductions are directly linked with European countries and substantial community spread contributed to the dispersion of the virus to smaller cities in the state country-side.

## MATERIAL AND METHODS

### Sampling and Molecular Detection

Samples were obtained from nasopharyngeal and oropharyngeal swabs from symptomatic patients from different ambulatory facilities in Pernambuco (Northeast Brazil). These samples are a part of the COVID-19 biorepository from the Aggeu Magalhães Institute (IAM), a Oswaldo Cruz Foundation (FIOCRUZ) unit. All standard operating procedures (SOPs) established by the World Health Organization guidelines were employed (2020c). Rigorous biosafety measures were employed as all samples were manipulated in the BSL-3 facility laboratory. RNA extractions were performed using the robotic platform using the Maxwell® 16 Viral Total Nucleic Acid Purification Kit (Promega, Wisconsin-USA), following the manufacturer’s protocol. The molecular detection was performed using the Kit Molecular Bio Manguinhos SARS-CoV-2 (E/RP): a single-step reaction for detecting the virus envelope gene (E) and the Ribonuclease P housekeeping control gene (RNAse P) [22].

### Genomic sequencing

Total RNA was used for single strand cDNA generation using Platus Transcriber RNase H- cDNA First Strand kit *(Sinapse inc)* following manufacturer’s instruction. cDNA generated was subjected to multiplex PCR reactions using Q5 High Fidelity Hot-Start DNA Polymerase (New England Biolabs) and a set of SARS-CoV-2 specific primers, designed by (https://www.protocols.io/view/ncov-2019-sequencing-protocol-bbmuik6w). Cycling conditions were: 98°C at 30 seconds, 98°C at 15 seconds, 62°C at 30 seconds and 65°C at 5 minutes during 35 cycles. Amplified PCR products were purified using AMPure XP Beads (Beckman Coulter) following standard protocol and quantified using the Qubit® dsDNA HS Assay Kits (Invitrogen) following the manufacturer’s instruction. Sequence libraries were prepared with Nextera XT Library Prep Kit (Illumina, San Diego, CA, USA) using 1.5 ng of PCR products following the manufacturer’s instructions. Sequencing was performed in the MiSeq (Illumina) machine using MiSeq Reagent kit V3 of 150 cycles employing a paired-end strategy.

### Genome Assembly and Annotation

Low quality raw sequencing reads and primers sequences were removed using Trimmomatic 0.36 with default parameters. Based on the knowledge that epidemic viruses sampled at short time frames does not accumulate a substantial amount of mutations, we performed a reference-based assembly strategy using the first published SARS-CoV-2 genome as reference (NC_045512.2) using Bowtie2 software [23] with default parameters. Following, we generated a .bed file using samtools 1.5 [24] and genomeCoverageBed from bedtools v 2.15.0 [25] keeping only position with > 5x of coverage. Lastly we used vcf-annotate (parameters --filter Qual=20/MinDP=100/SnpGap=20) and vcf-consensus from vcftools v 0.1.13 [26] to generate the final consensus sequences.

The N regions and coverage values for each genome were plotted using karyoploteR [27] and the annotation was performed with VAPiD [28], using the NC_045512.2 genome as reference. All in house scripts used in the following sections are deposited on https://github.com/dezordi/SARS-CoV-2_tools.

### Evolutionary Analysis

Nineteen thousand nine hundred and one public genomes of SARS-CoV-2 were retrieved from GISAID (https://www.gisaid.org/ - accessed in 23 May 2020), this number represents only SARS-CoV-2 genomes identified in human samples tagged as complete and sequenced with high coverage. Sequences with less than 29400 bp were removed with fasta_cleaner.py resulting in 16,645 genomes. These genomes and the 38 ones sequenced in our study were aligned with the reference genome NC_045512.2 using MAFFT add v7.310 [29] with the --keep-length parameter. The resulted alignment was edited in different ways to generate three datasets: I - The 3’ and 5’ ends were removed according to [30]; II - The gaps and homoplasic sites were masked according to (https://virological.org/t/issues-with-sars-cov-2-sequencing-data/473) with algn_mask.py and III - Only the gaps and nucleotide positions with degenerated IUPAC bases were masked. Each alignment was submitted to a clustering step using cd-hit-est [31] to remove redundancy with cluster_gisaid.py, where sequences with 100% of identity from the same country at the nucleotide level were clustered and only one representative genome were selected for further analyses. Such subsampling strategy was necessary since the sequencing effort has been much more intense in some countries than others. Still, after this first redundancy removal more than 3000 genomes remained for the USA and England. Then a second round of redundancy removal was applied using a lower identity threshold (99.7%) only for some countries (see Supplementary Material 1 - Sheet 1) in order to sample a similar number of genomes available for the majority of the remaining countries (ranging from 153 to 194). It is important to note that we kept all the information about clustered sequences and added them back to the alignment if some of our sequences grouped with high branch support to one of the representative sequences from the clusters in order to reduce any bias to the final phylogenetic reconstruction. Finally, each alignment dataset was visualized and checked with Aliview [32] and these 3 alignments were submitted to the following steps.

The SARS-CoV-2 lineages were assigned with pangolin (https://pangolin.cog-uk.io/) and the phylogenetic trees were reconstructed with IQ-TREE [33] using 1000 replicates of the bootstrap ultrafast method [34]. The evolutionary models were selected with the ModelFinder [35] for each dataset. The best model of substitution, log likelihood values for each datasets is available on (Supplementary Material 1 - Sheet 2). iTOL [36] was used to annotate the phylogenetic trees, the annotation files were generated with itol_annot.py and pangolin_annot.py. Tree branches were colored by continent and tip color ranges were colored by pangolin lineage, the root was set between lineages A and B, as proposed by [30]. After each tree reconstruction and annotation, the dataset that generates the tree with best log-likelihood value, and a topology that converges with pangolin annotation was selected and subsets of each clade containing IAM genomes were created to rerun phylogenetic trees for each clade (Supplementary Material 1 - Sheet 2) with the same parameters of the previous tree. After tree reconstructions, genomes from GISAID that are present on highly-supported clades (branch support ≥ 90) with IAM genomes were separated to evaluate the SNPs supporting each cluster. Information about collection date and travel history used in subsequent analysis can be found in Supplementary Material 1 - Sheet 3.

For the Bayesian analysis, datasets with SARS-CoV-2 genomes were sampled by week with gisaid_sampler.py and only sequences showing complete collection dates (year-moth-day) were kept. One dataset was constructed comprising the lineages B + B1 from pangolin including all SARS-CoV-2 genomes sequenced in this study. After the ML reconstruction with IQ-TREE the tree was evaluated in Tempest 1.5.3 [37] to check the root-to-tip temporal signal. Outlier sequences were removed before the phylodynamics analysis performed in BEAST 1.10.4 [38]. Bayesian phylodynamic analyses were performed using either a strict clock applying a fixed mean clock rate of 0.8×10^-3^ as used in other studies [13,39] or an uncorrelated lognormal relaxed clock model. The analyses were performed either using a constant, coalescent exponential growth or Skyline demographic model as tree priors and GTR+F+I as nucleotide substitution model. The molecular clock and demographic model were evaluated based on likelihood using Path Sampling/Stepping-stone sampling. The final analysis was based on three independent runs of 100 million MCM generations sampling in every 10.000 steps using the uncorrelated lognormal relaxed clock and coalescent exponential growth demographic model. The run convergence was evaluated with Tracer 1.7 and all parameters showed ESS values > 200. The generated trees were combined using LogCombiner applying a burn-in of 25% and the Maximum clade credibility tree was obtained using the treeannotator. The time-scaled trees were visualized on Figtree 1.4.4.

### Single Nucleotide Polymorphism analysis

Single Nucleotide Polymorphism (SNPs) were evaluated with the snp-sites tool [40], using as input the alignments containing the genomes generated in this study and the NC_045512.2 genome as reference. The positions with SNPs were retrieved with BCFtools [41]. The SNPs positions were crossed with the depth of genome assembly and nucleotide diversity per position accessed with bam-readcount tool (https://github.com/genome/bam-readcount), the outputs from bcftools and bam-readcount were crossed with snp_div.py to access the metrics and nucleotide diversity of SNPs by genomic region (Supplementary Material 4 and 5). Owing to the importance of Spike protein in SARS-CoV-2 biology, Single Amino acid Polymorphisms (SAPs) and regions were deletions were found in other SARS-CoV-2 genomes [42] were carefully analyzed using Aliview and karyoploteR (http://bioconductor.org/packages/release/bioc/html/karvoploteR.html).

### Epidemiological data

SARS-CoV-2 confirmed and deaths cases from Brazil and Pernambuco state were obtained from the coronavirus website from the brazilian Ministry of Health (https://covid.saude.gov.br/). We collected the number of cases and deaths per day and per epidemiological week since the first case reported in the state (March 12th, 2020). Plots were performed using the ggplot2 package of the R statistical language (https://www.r-proiect.org/).

## RESULTS AND DISCUSSION

### Epidemiological data from Pernambuco state

The thirty-eight samples processed for whole genome amplification and sequencing were obtained from eight municipalities of the Pernambuco state (Figure 1A). The first confirmed SARS-CoV-2 infection in Pernambuco was reported in the second week of March (12th March 2020), in the 11th epidemiological week (Figure 1B), only sixteen days after the first confirmed case in Brazil (25th February 2020) (Figure 1B - top-left panel). In the 21st epidemiological week, the Pernambuco state reached the peak of the pandemics reporting 8298 new cases, averaging 1185 new cases per day (Figure 1 - top-right panel). An increasing number of deaths followed, in which the largest number occurred at the twenty-first epidemiological with 683 deaths and an average of 97.57 deaths per day (Figure 1 - lower panel). The thirty-eight samples selected for SARS-CoV-2 whole genome sequencing were collected at the 15th (32 samples) and 16th (six samples) epidemiological weeks (first and second weeks of April - blue bars in Figure 1B - lower panel) representing the beginning of the SARS-CoV-2 spread at the Pernambuco state.

**Figure 1.**
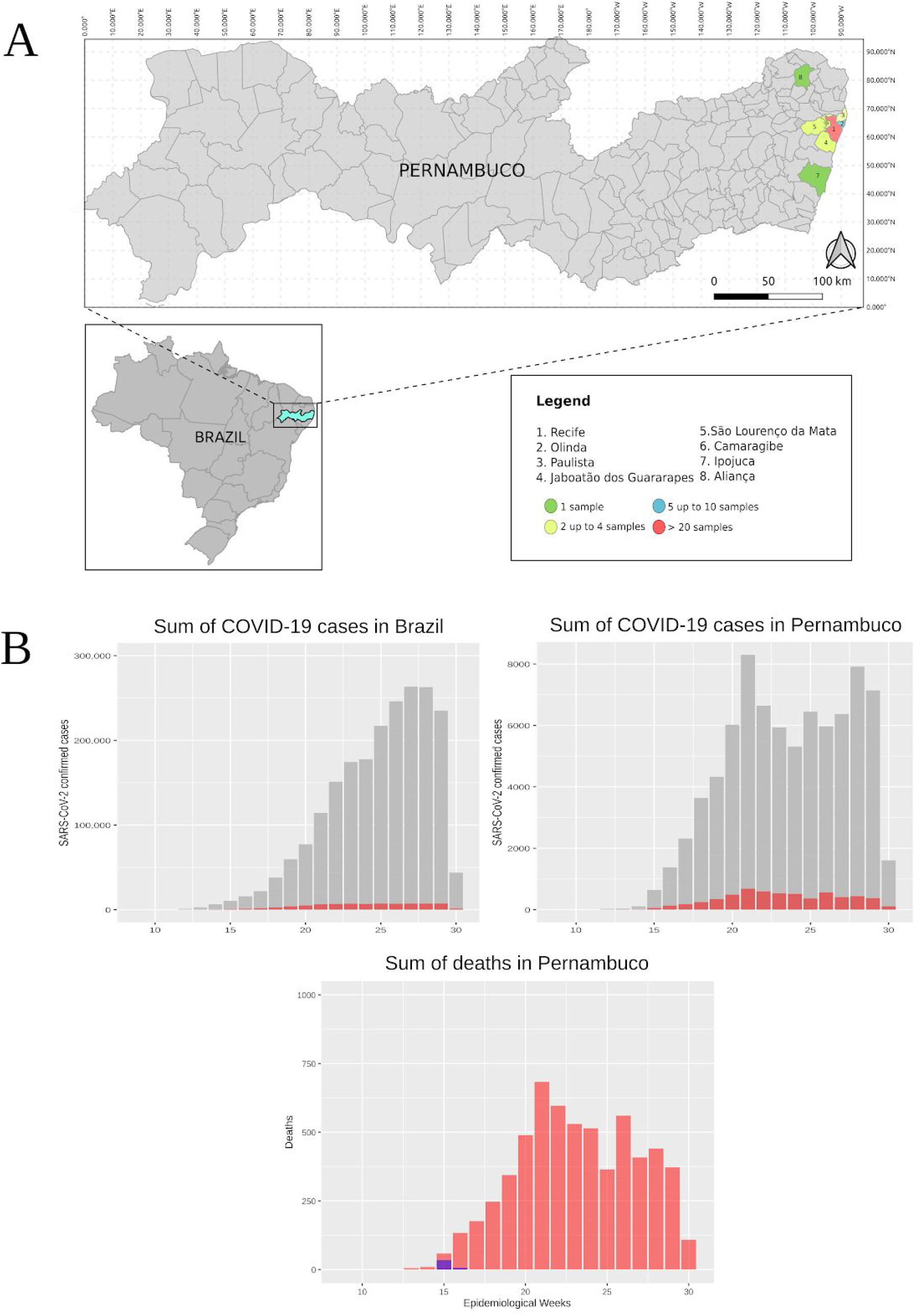
Sample distribution over the Pernambuco state municipalities and epidemiological curves of SARS-CoV-2 confirmed patients from Brazil and Pernambuco per epidemiological week. A - Map showing the sample distribution by Pernambuco municipality. B - SARS-CoV-2 epidemiological data from Brazil - Top-left panel and Pernambuco state - Top-right and lower panels. Number of confirmed SARS-CoV-2 cases (grey) and deaths (red) per epidemiological week. Number of genomes sequence (blue bars) obtained in this study. Epidemiological week 10 refers to the first week of March 2020 and the first official case reported in Pernambuco on March 12th 2020.

### Genome sequencing and variability

Thirty-eight SARS-CoV-2 genomes were obtained with a coverage breadth ranging from 93.92 to 99.92. All genomes were considered of high quality following the GISAID criteria (showing coverage higher than 95%) except for sample IAM138 (Table 1). We also obtained a very high average coverage depth of 2354x with a standard deviation of 868x (Table 1).

**Table 1.** Information of human samples processed for SARS-CoV-2 genome sequencing.

| Sample name | Collection date | Municipality | SARS-CoV-2 qRT-PCR Ct value | Genome coverage breadth | Genome average coverage depth | SARS-CoV-2 lineages assigned by Pangolin* |
| --- | --- | --- | --- | --- | --- | --- |
| IAM08 | 2020-04-07 | Recife | 16.189 | 99.47 | 2894.98 | B.1.1 |
| IAM10 | 2020-04-07 | Recife | 16.682 | 99.48 | 2846.24 | B.1.1 |
| IAM16 | 2020-04-07 | Recife | 17.36 | 94.93 | 1531.11 | B.1.1 |
| IAM17 | 2020-04-07 | Recife | 16.8 | 96.97 | 3121.49 | B.1.1 |
| IAM18 | 2020-04-07 | Recife | 14.88 | 97.09 | 1648.15 | B.1.1 |
| IAM19 | 2020-04-07 | Recife | 16.14 | 94.76 | 1706.99 | B.1 |
| IAM29 | 2020-04-08 | Olinda | 18.352 | 99.17 | 2867.59 | B.1.1 |
| IAM30 | 2020-04-08 | Ipojuca | 18.166 | 99.44 | 2947.33 | B.1.1 |
| IAM39 | 2020-04-08 | Recife | 14.91 | 98.59 | 3342.17 | B.1.1 |
| IAM48 | 2020-04-08 | Recife | 17.465 | 98.86 | 2081.51 | B.1.1 |
| IAM67 | 2020-04-08 | Recife | 14.456 | 97.86 | 2702.48 | B.1.1 |
| IAM84 | 2020-04-08 | Recife | 13.891 | 99.89 | 3185.84 | B.1.1 |
| IAM87 | 2020-04-08 | São Lourenço da Mata | 15.517 | 98.84 | 2667.67 | B.1.1 |
| IAM89 | 2020-04-07 | Camaragibe | 12.367 | 99.5 | 3348.85 | B.1.1 |
| IAM103 | 2020-04-09 | Recife | 11.785 | 99.3 | 2568.56 | B.1.1 |
| IAM138 | 2020-04-09 | Recife | 16.751 | 89.59 | 740.25 | B |
| IAM139 | 2020-04-09 | Recife | 15.42 | 96.24 | 1971.05 | B.1.1 |
| IAM158 | 2020-04-09 | Recife | 19.626 | 96.73 | 2942.74 | B.1.1 |
| IAM167 | 2020-04-09 | Recife | 17.18 | 96.05 | 1659.19 | B.1.1 |
| IAM171 | 2020-04-09 | Recife | 18.39 | 96.45 | 1339.42 | B.1.1 |
| IAM209 | 2020-04-09 | Recife | 15.978 | 97.15 | 1854.91 | B.1.1 |
| IAM211 | 2020-04-09 | Recife | 16.097 | 95.14 | 2064.34 | B.1.1 |
| IAM212 | 2020-04-09 | Recife | 15.928 | 97.24 | 2218.94 | B.1.1 |
| IAM215 | 2020-04-06 | Olinda | 17.33 | 97.88 | 3669.32 | B.1.1 |
| IAM220 | 2020-04-09 | Recife | 18.698 | 96.95 | 3499.37 | B.1.1 |
| IAM221 | 2020-04-08 | Recife | 15.615 | 99.18 | 3157.48 | B.1.1 |
| IAM226 | 2020-04-07 | Jaboatão dos Guararapes | 14.879 | 98.17 | 2011.33 | B.1.1 |
| IAM230 | 2020-04-09 | Recife | 16.878 | 95.1 | 2274.38 | B.1.1 |
| IAM235 | 2020-04-09 | Recife | 18.442 | 95.8 | 2204.80 | B.1.1 |
| IAM238 | 2020-04-09 | Recife | 17.844 | 97.78 | 4397.48 | B.1.1 |
| IAM248 | 2020-04-09 | Recife | 19.652 | 93.77 | 1494.11 | B.1.1 |
| IAM273 | 2020-04-08 | Recife | 17.761 | 97.08 | 2135.29 | B.1.1 |
| IAM291 | 2020-04-13 | Recife | 20.235 | 94.32 | 1508.67 | B.1.1 |
| IAM305 | 2020-04-13 | Camaragibe | 20.278 | 96.11 | 3152.63 | B.1.1 |
| IAM307 | 2020-04-13 | Aliança | 21.155 | 94.52 | 1415.27 | B.1.1 |
| IAM311 | 2020-04-13 | Recife | 17.311 | 98.02 | 1131.23 | B.1.1 |
| IAM355 | 2020-04-13 | Olinda | 18.149 | 97.38 | 2771.64 | B.1.1 |
| IAM356 | 2020-04-13 | Paulista | 18 | 94.14 | 382.87 | B.1.1 |

\* Lineages were assigned with pangolin (https://pangolin.cog-uk.io/) updated on 03 June 2020.

Coverage depth was mostly uniform with few amplicons showing a much higher depth and few systematic gaps between the 7th and 8th kb position from the ORF1ab, the ORF that generate all non-structural proteins of SARS-CoV-2, and between 20-21kb of the Spike protein ORF (S), the outermost protein in the coronavirus crown responsible for the cell surface receptor binding [43] (Figure 2 and Supplementary Material 2. These systematic gaps are probably related to primer competition during the PCR reaction, which probably lead to a higher abundance of some amplicons in detriment of others. It can be clearly seen in the coverage depth plot for each genome in Supplementary Material 2. On the other hand, at least for the Spike protein deletions, Liu et al 2020 [27] found recurrent deletions in the coding region that may restrict late phase viral replication both in clinical and in vitro isolated viral strains [42]. In order to investigate if these natural occurring deletions could be responsible for such gap patterns seen in the spike protein of the genomes sequenced here we manually checked the alignment of all genomes from Pernambuco and probed our raw sequencing paired-end data. We could not detect any evidence of deletion in the Spike protein region (NSPRRAR) or (QTQTN) (Supplementary Material 3) identified by Liu et al. 2020 suggesting that the gap regions found in the genomes sequenced here were likely a result of primer competition and the low amount of sequenced amplicons corresponding to those regions.

**Figure 2.**
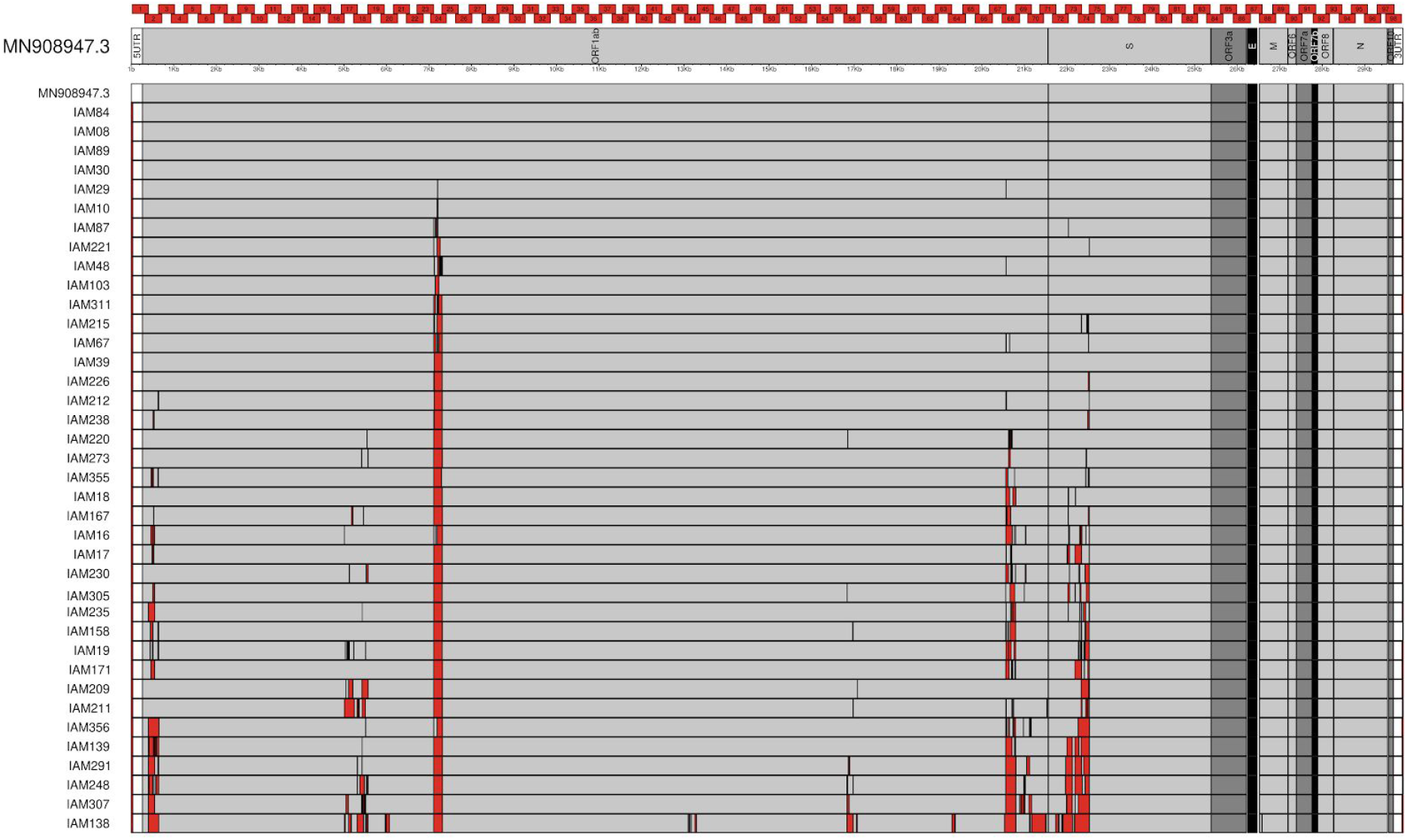
SARS-CoV-2 genomic map of the reference strain Wuhan-1 (MN908947) and the genomes obtained in this study. Thirty-eighth SARS-CoV-2 genomes sequenced using whole genome amplification and a paired-end sequencing strategy (2×75 bp). Red rectangles above the Wuhan-1 reference genome are the overlapping amplicons obtained with the NetworkArtic V3 primer sets (https://artic.network/resources/ncov/ncov-amplicon-v3.pdf) while red spaces in the sequenced genomes correspond to non sequenced regions.

Regarding amino acid mutations, new evidence recently emerged showing amino acid change in the Spike protein. An amino acid change (D614G) is of particular interest since G614 lineages have consistently replaced well established D614 strains and could confer a fitness advantage of the former strains leading to a higher viral load in infected patients and higher mortality rate [44-46] although it is not clear yet if this mutation has any impact on virus transmissibility and covid-19 progression and outcome [47]. Besides, this amino acid change is almost always accompanied by three other mutations: a non coding nucleotide change (C-to-T) in the 5'UTR, a synonymous nucleotide mutation (C-to-T) at position 3,037 and a non-synonymous mutation (C-to-T) at position 14,408 that generated an amino acid change in the RNA-dependent RNA polymerase (RdRp P323L). All genomes sequenced from the Pernambuco state showed the G614 amino acid change and the three accompanying nucleotide mutations described above, except the IAM138 strain which has all four positions identical to Wuhan-1 reference sequence (Figure 3. Supplementary Material 4). Recent experimental evidence, in different human cell lines, confirmed that C614G is a key amino acid change that increases SARS-CoV-2 infectivity and when associated with I472V may increase the resistance to neutralizing antibodies [44]. Interestingly, two hundred and two SARS-CoV-2 genomes available from Brazil have been screened for such mutations (up to 30th July 2020) and 190 of those contain the D614G mutation (https://cov.lanl.gov/apps/covid-19/map/). Such data associated with the results from our study suggests that G614 also replaced D614 strains in a similar way as it happened in other countries or it is a result of a founder effect based on the importation of primarily G614 variants to Brazil. However, additional genomic data will be necessary to evaluate those two hypotheses.

The full set of nucleotide changes found in the genomes sequenced in this study can be found in Supplementary Material 1 - Sheet 4 and 5)

**Figure 3.**
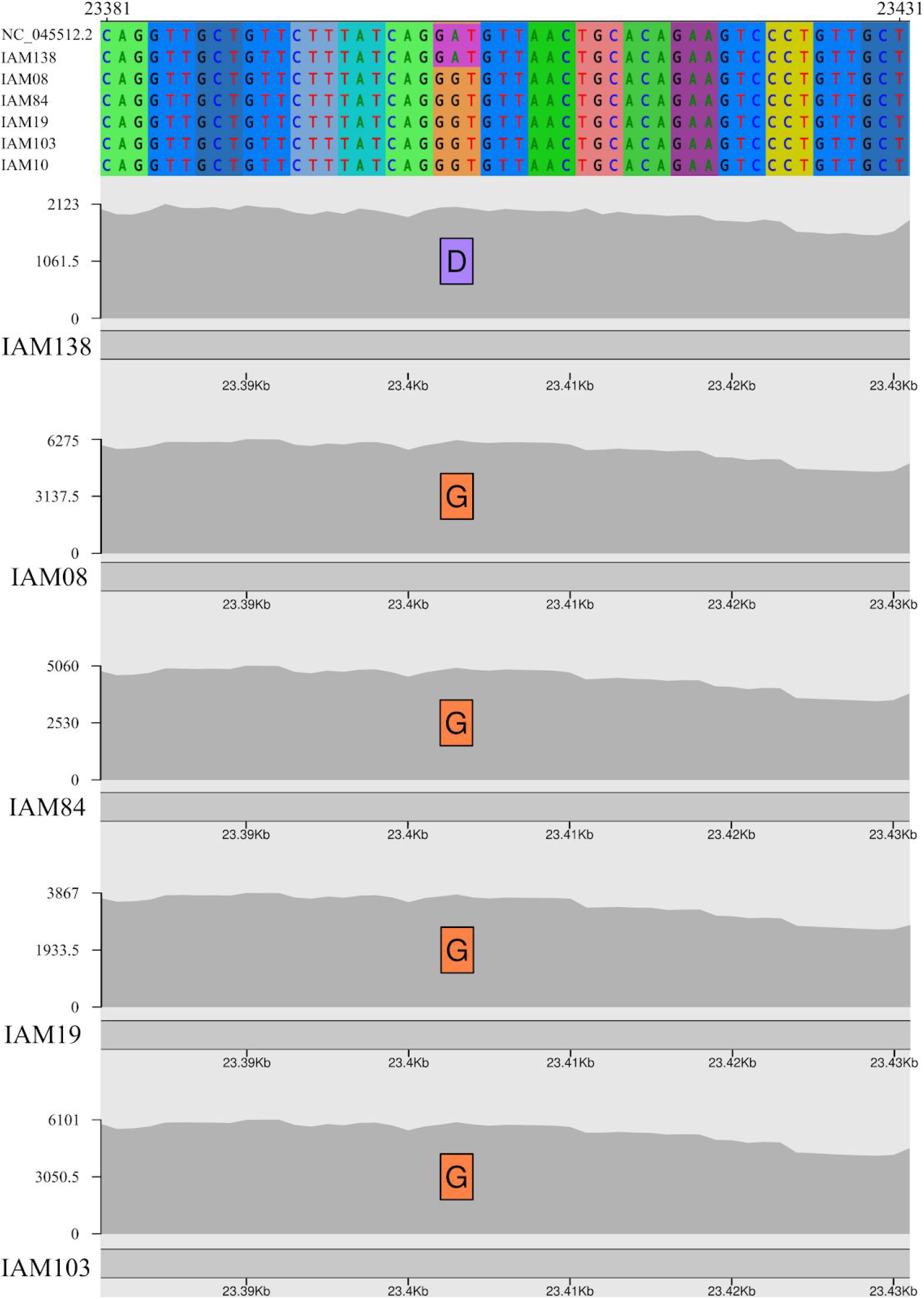
Spike protein amino acid change D614G found in the genomes sequenced in this study. Codon alignment of SARS-CoV-2 Wuhan-1 Spike coding region showing the reference D amino acid in the IAM138 genome and the G amino acid in other four representative genomes sequenced in this study. x axis represents Wuhan-1 genomic coordinates and the y axis the coverage depth at each position.

### Phylogenetic clustering of Pernambuco SARS-CoV-2 strains

Comprehensive analysis with thousands of genomes have identified two main SARS-CoV-2 lineages called A and B, which emerged during the beginning of the pandemic in the Hubei province - China [30]. Besides, a subdivision of these lineages was also proposed with 5 and 9 sub lineages belonging to A and B lineages, respectively. Both A and B major lineages spread worldwide, but ongoing studies have been showing that the B lineage spread and replaced the lineage A in several different countries [45]. One clear example of that was the B1 lineage, that was responsible for the Italian outbreak [48,49], and it later spread to other European countries and several countries in the Americas including Brazil [20]. Up to now, three studies investigated the SARS-CoV-2 lineages imported to Brazil and its further spread into the country. The largest study sequenced 427 genomes and analyzed 490 genomes from 21 brazilian states detecting that only 5 strains belong to the lineage A while the remaining 485 belong to the B lineage [20]. These authors estimated that more than 100 international introductions of the virus occurred in Brazil. Resende et al 2020 sequenced 95 genomes from 10 Brazilian states characterizing six SARS-CoV-2 lineages (A.2, B.1, B.1.1, B.2.1, B.2.2 and B.6). In agreement with the above, the majority of the strains were classified as clade B.1 (95%) and 92% of those belong to the sub-clade B.1.1 [19]. Lastly, Xavier et al 2020 sequenced the genome of 40 strains of SARS-CoV-2 from Minas Gerais state where 85% of these belong to the B lineage in which most of those fall within B1.1 and only one genome belong to the A lineage [18]

In order to evaluate which lineages belong the genomes sequenced in this study, we performed the lineages and sublineages assignment with Pangolin following Rambout et al 2020 dynamic nomenclature for SARS-CoV-2 and plotted it onto a ML phylogenetic tree reconstructed with 5297 SARS-CoV-2 genomes (Figure 4). All thirty-eight viral genomes obtained from Pernambuco belong to the B lineage, one strain (IAM138) was basal to all B lineages but with low branch support (Figure 4A). This same genome presented the lowest genome coverage breadth (89%) (Table 1) and such clustering could be related to the lower amount of sequenced synapomorphic single nucleotide polymorphism (SNPs). However, we found several nucleotide and amino acid polymorphisms that are characteristic of early diverging SARS-CoV-2 lineages such as the reference Wuhan-1 genome sequencing (see **Genome sequencing and variability** section above and **Figure 3**) suggesting that this lineage is likely a representative of the early divergent SARS-CoV-2 lineages emerged from China.

A second strain (IAM19) was assigned to the B.1 lineage and grouped with high branch support with two SARS-CoV-2 genomes from France (Figure 4 B). The remaining thirty-six strains grouped in the B1.1 lineage (Table 1 and Figure 4 C), but 10 were placed directly at a polytomic branch showing no specific clustering with other samples. The proofreading correction of the RNA polymerase of SARS-CoV-2 genome results in a low mutation rate of the SARS-CoV-2 compared to other RNA viruses [50] and such low variability of densely sequenced genomes from a short period of time likely hindered the positioning of those samples since there is a low number of informative SNPs available for a confident phylogenetic clustering from the early pandemic phase. In addition, IAM311 sample clustered with a genome from Belgium with posterior probability of 0.83 (**Figure 4C**). On the other hand, at least twenty-three strains grouped between them in clades containing only Pernambuco samples with a high posterior probability branch support (over 70). It clearly reveals virus transmission chains between neighborhoods and municipalities of the Pernambuco state corroborating the rapid community transmission within the state (**Figure 4 C**). In addition, two samples (IAM67 and IAM356) grouped with high branch support with a sample from Alagoas, a southern neighbouring state (**Figure 4 C**) and IAM307 grouped with several genomes from South America having as a sister group of SARS-CoV-2 genomes from European countries, both highly-supported (**Figure 4 C**).

Time-resolved bayesian tree reconstructions confirmed all clades (Figure 4) and the polytomic positioning of several of the genomes investigated here as shown in the ML tree including the basal positioning of IAM138 samples although with no branch support (Figure 5). Besides, the estimated most recent common ancestors (tMRCA) of specific clades are in agreement to the emergence of the virus around December 2019. The IAM19 genome clustered with two samples from France with an estimated the more recent common ancestor (tMRCA) between late February and mid March (Table 2) suggesting that such viral strain correspond to one of the first importation cases to the state in agreement with the first official case reported (March 12). Moreover, all other sequenced strains belonging to the B1.1 lineage that were clustered in highly supported clades showed overlapping tMRCA between mid March and Mid April (**Table 2**) supporting that these lineages that emerged slightly later were the most successful ones spreading in the state. (**Figure 5**).

Overall, our results showed two new international introduction events of the SARS-CoV-2 from Europe, spread of the virus between brazilian states and the community transmission in the Pernambuco state. Besides, the lineage assignment further supports that the B lineage is prevailing through community spread in Brazil in line with other SARS-CoV-2 studies.

**Figure 4.**
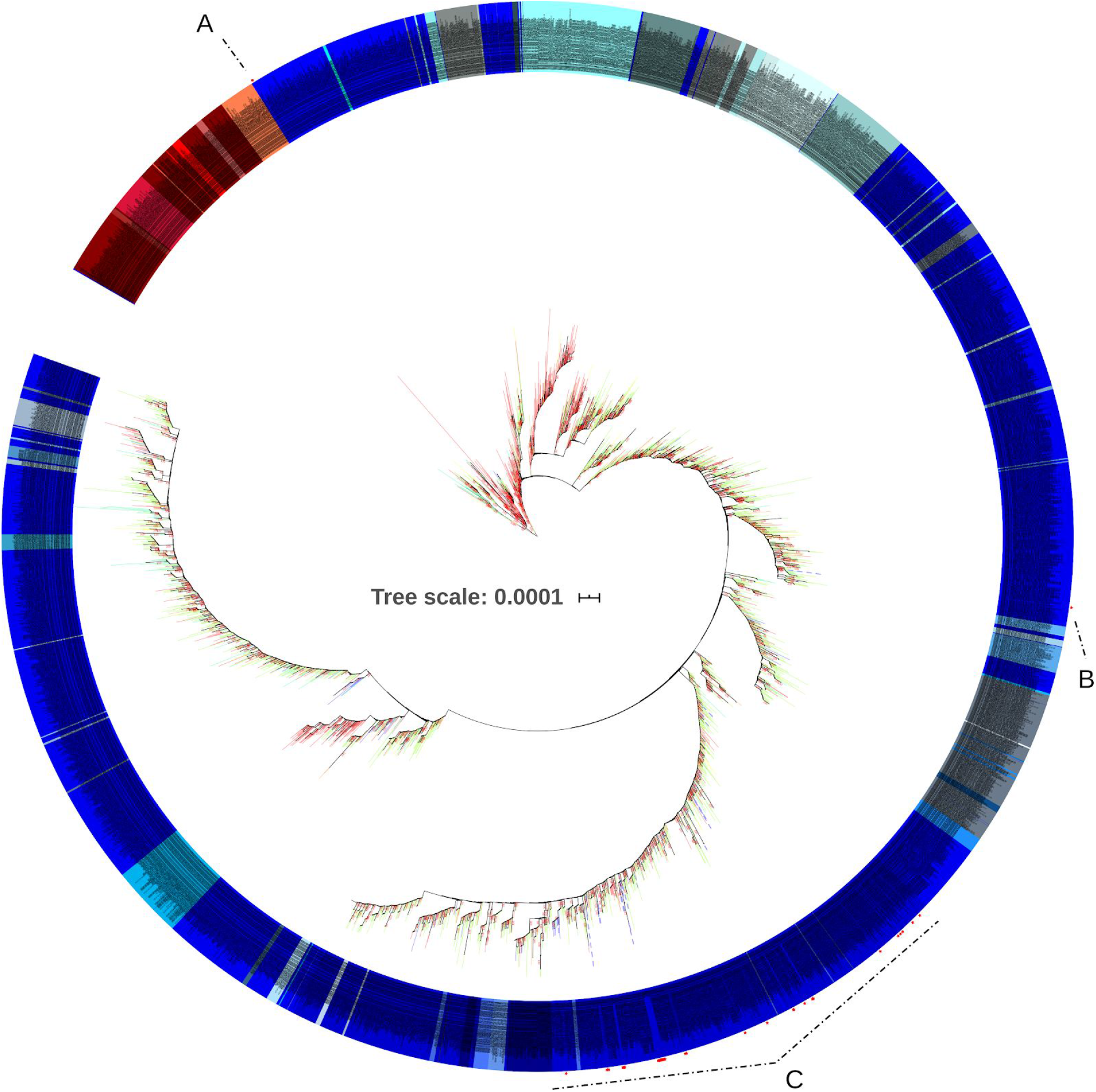
Maximum likelihood phylogenetic tree using 5259 genomes available from GISAID plus 38 genomes generated in this study (red stars) rooted between group A and B. Pangolin lineage assignments are denoted by the tip colors: reddish are A lineages and bluish are B lineages. Branch colors follows continent of samples origin: Red - Asia; Orange - Africa; Yellow - Australia and Zealandia; Green- Europe; Pink - North America; Blue - Central America; Purple - South America. Full interactive tree can be found at https://itol.embl.de/tree/45462241443551591453841.

**Figure 5.**
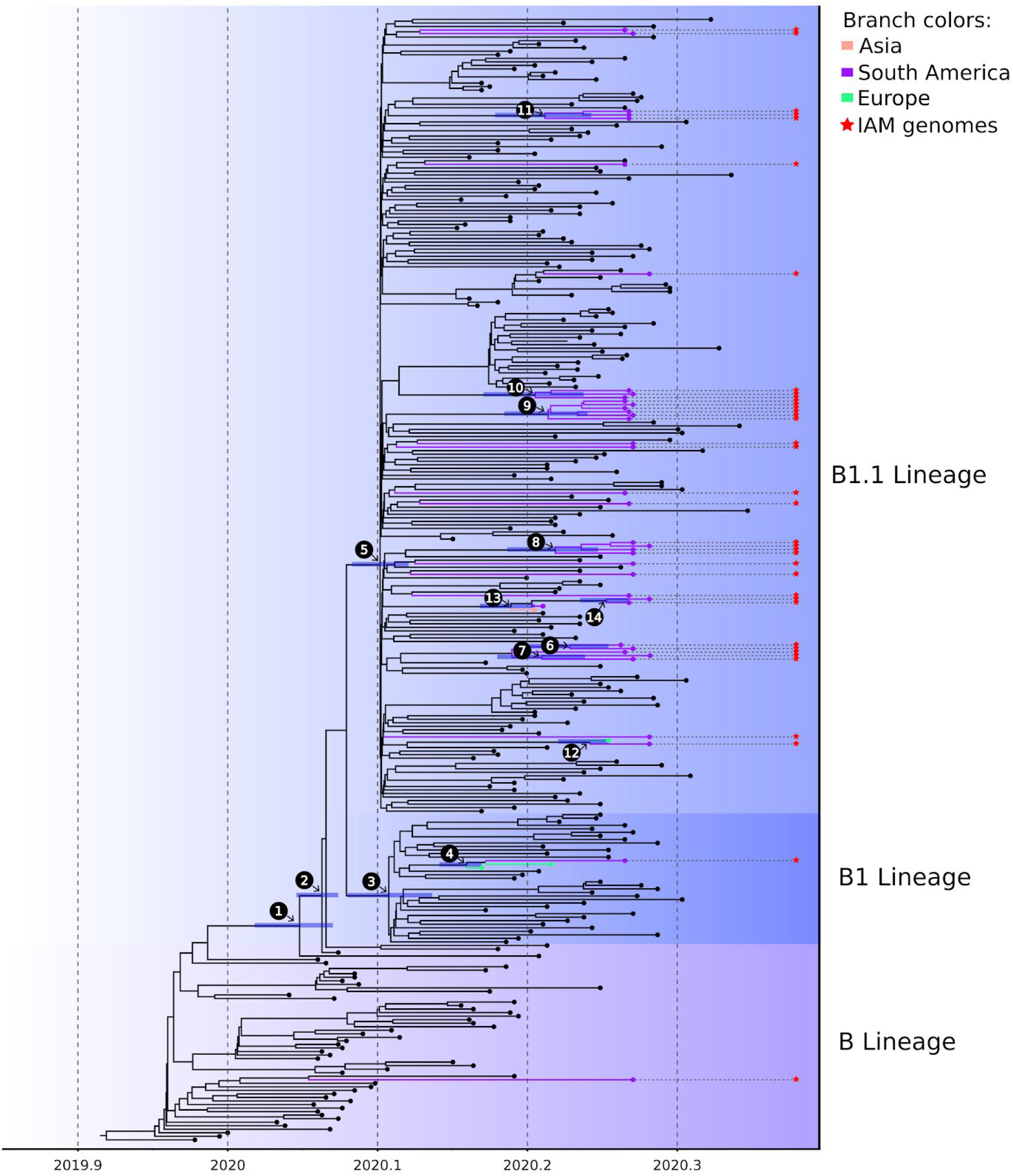
Time-resolved tree of the B lineage reconstructed with a Bayesian framework. Numbers above the branches are specific clades showing posterior probability higher than 80 depicted in Table 2 and horizontal bar represents the HPD95% credible interval of the estimated tMRCA. Branch colors follow continent order as in Figure 4, but only highly supported clades having the sequences obtained in this study were colored. Red stars show the sequenced IAM genomes.

**Table 2.** Predicted origin intervals of clades from phylodynamic analysis.

| Node number* |  | HPD95% time interval |
| --- | --- | --- |
| 1 |  | Early to late January |
| 2 |  | Early to late January |
| 3 | B1 lineage ancestral | Late January to late February |
| 4 | Inferred international importation from Europe | Late February to mid March |
| 5 | B1.1 lineage ancestral | Early February to late February |
| 6 | Local transmission | Mid March to mid April |
| 7 | Local transmission | Mid March to late March |
| 8 | Local transmission | Mid March to early April |
| 9 | Local transmission | Mid March to early March |
| 10 | Local transmission | Mid March to late March |
| 11 | Local transmission | Mid March to late March |
| 12 | Inferred international importation from Europe | Late March to mid April |
| 13 | Local transmission | Mid March to late March |
| 14 | Local transmission | Late March to mid April |

\*Nodes shown in Figure 5.

## CONCLUSION

The genomic analysis of 38 SARS-CoV-2 genomes from the beginning of the epidemic at Pernambuco state - Brazil revealed at least two independent international importation events of SARS-CoV-2 strains from Europe occurring from late February to mid March and a more recent one from late March to mid April which is in line with studies focusing on other Brazilian states. Moreover, we also found evidence of comunitary spread of SARS-CoV-2 among Recife city neighborhoods and other municipalities. Interestingly, all genomes belong to the B lineage, with a higher prevalence of B1.1 lineage, which was shown to be the most prevalent currently circulating in Brazil. All except one genome have the G614 amino acid change which has been suggested to increase the viral fitness allowing to reach a higher viral load in an experimental setting and likely in human patients. G614 strains have replaced D614 strains in several regions around the globe where the second was first established. The high prevalence of G614 strains in our dataset suggests two possible explanations: I - most strains that entered Brazil belonged to G614 strains or D614 and G614 were equally imported into Brazil but G614 became prevalent as it occured in European and North American countries. Continued genomic-based surveillance of SARS-CoV-2 in a much broader set of samples is needed in order to access the viral mutational spectra and provide data to tease apart those hypotheses that will likely impact the control measures set to curb the epidemic.

## Data Availability

All data can be accessed in the GISAID databank (https://www.gisaid.org/)

## DATA AVAILABILITY

All genomes generated in this study are deposited on GISAID under the accessions EPI_ISL_500460-500486 and EPI_ISL_500865-500875.

## ACKNOWLEDGMENT

We thank the many brave researchers around the globe for their effort to generate and make readily available the SARS-CoV-2 genomic data in GISAID and other databases. We also thank the LACEN-PE whole team for providing the samples to sequence the SARS-CoV-2 genomes, the Technological Platform Core and the Bioinformatic Core of the Aggeu Magalhaes Institute for the support with their research facilities. This project was supported by the National Council for Scientific and Technological Development by the productivity research fellowship level 2 for Wallau GL (303902/2019-1).

## AUTHOR CONTRIBUTIONS

LCAB, SPBF, CFJA and GLW conceived and planned the study. RHS. PMSL, BASA, DGAC, SCGA, JJFM, MJCO, LVBL and PROH conducted swab collections and sample processing. MHSP, DRDG, CD, MFB, FZD, LCM, LK, EH, AFS, SJRS, KGSS, BSLFS, DLC, CEC, AMN, CTAS, RPGM, MALS, MSB, WRCN, RMLA performed laboratory experiments. MHSP, DRDG, CD, MFB, FZD, AFS, LCM, LRSV, AMR, CFJA and GLW performed data analysis. MHSP, FZD and GLW wrote the manuscript. All authors reviewed the final manuscript.

## CONFLICTS OF INTEREST

The authors declare no conflict of interest.

**KARYOPLOTER**: karyoploteR. http://bioconductor.org/packaaes/release/bioc/html/karvoploteR.html (Accessed on 03 June 2020).

**SARS-COV-TOOLS**: SARS-CoV-2 tools. https://github.com/dezordi/SARS-CoV-2_tools (Accessed on 03 June 2020).

**VIROLOGICAL**: Virological.org. https://virological.org/t/issues-with-sars-cov-2-sequencing-data/473 (Accessed on 03 June 2020).

**BCF-TOOLS**: BCFtools. https://github.com/samtools/bcftools (Accessed on 03 June 2020).

**BAM-READCOUNT**: bam-readcount. https://github.com/genome/bam-readcount (Accessed on 03 June 2020).

